# Domestic Gender-Based Violence and Sexual and Reproductive Health-Seeking Behavior Among Women in Lusaaze Informal Settlement, Kampala, Uganda

**DOI:** 10.64898/2026.07.05.26355955

**Authors:** Kavuma Sharif, Nsungwa Elizabeth

**Affiliations:** Kampala Hospital Limited - Kampala, Uganda; Victoria University-Kampala, Uganda

## Abstract

**Background:** Domestic gender-based violence (DGBV) can undermine women’s sexual and reproductive health (SRH) and reproductive autonomy. However, community-level evidence examining multiple forms of violence and their relationship with SRH-seeking behaviour in Uganda’s informal urban settlements remains limited. This study assessed the prevalence of DGBV, associated factors and its relationship with SRH-seeking behaviour among women in Lusaaze, Kampala.

**Methods:** A quantitative, community-based analytical cross-sectional study was conducted among 383 women aged 15–49 years. Data from 377 eligible participants were analysed using descriptive statistics and modified Poisson regression with robust standard errors to estimate adjusted prevalence ratios (aPRs) and 95% confidence intervals (CIs).

**Results:** Among 374 women with complete violence data, 72.5% (95% CI 67.7–76.7) reported at least one form of DGBV, while 49.7% experienced all four forms. Emotional violence was most prevalent (65.2%), followed by physical (62.8%), sexual (59.9%) and economic violence (56.7%). DGBV was not significantly associated with HIV testing or antenatal care attendance. However, it was associated with current contraceptive use (aPR 1.11, 95% CI 1.01 – 1.22) and partner interference with family planning (aPR 3.36, 95% CI: 1.25 – 9.06).

**Conclusion:** DGBV was highly prevalent and frequently involved multiple forms of violence. Despite high contraceptive use and antenatal care attendance, reproductive autonomy remained constrained by partner interference. Integrating confidential, survivor-centred violence screening and reproductive autonomy support into SRH services is essential.

## Background

Domestic gender-based violence (DGBV), encompassing physical, sexual, emotional and economic violence within intimate relationships, is a major public health and human rights concern [1,2]. Data from the World Health Organization (WHO) suggests that approximately one in three women globally (nearly 840 million women aged 15 years and above) has experienced physical and/or sexual violence from intimate partners or non-partners in her lifetime, which has remained roughly constant over the last 20 years. While physical and sexual abuse have historically garnered the most clinical and policy attention, abuse in the form of emotion control and economic control are likely to co-occur with physical violence and, together, render poly-victimisation-which may lead to enhanced mental health outcomes and reduced women’s ability to earn a living from the household [2,3].

In addition to the physical injuries, domestic gender-based violence (DGBV) has detrimental and long-term effects on women’s sexual and reproductive health (SRH). There is strong, consistent evidence that women experiencing DGBV are at increased risk of unwanted pregnancy, unsafe abortion, STIs, HIV and adverse pregnancy outcomes [4,5]. DGBV may also be a structural and interpersonal barrier to accessing health care services. Threatening partner behavior, fear of retribution, financial constraints and internalized stigma may diminish women’s confidence to negotiate contraceptive use, to access appropriate ANC, to attend regular HIV testing sessions, or to obtain immediate treatment of STI symptoms [4–6].

DGBV and negative sexual and reproductive health (SRH) outcomes are especially intertwined in Sub-Saharan Africa in settings where male supremacy, economic instability and infectious disease and endemic violence are prevalent [7]. Meta-analyses at the regional level have demonstrated that lifetime prevalence of intimate partner violence in Sub-Saharan Africa is higher than the global estimate, at 36–44%, for ever partnered women. Ugandan women continue to experience violence towards women in spite of progressive legal frameworks such as the Domestic Violence Act (2010) and the National Policy on Elimination of Gender-Based Violence [7,8]. In 2022, the Ugandan Demographic and Health Survey (UDHS) reported that 43.7% of all women (aged 15-49) ever in a union experienced physical violence and 16.7% (both physical and sexual violence combined) experienced sexual violence from their current or most recent intimate partner. Prevalence of SRH service gaps such as high levels of unmet need for modern contraception, inadequate attendance to ANC, and postponement of STI/HIV services are likely leading to preventable maternal morbidity and transmission of HIV among women of reproductive age in Uganda [9,10].

In Uganda, the rapid growth of informal urban settlements lacking essential resources has increased community-level socioeconomic stresses that may influence both household violent interactions and access to SRH services for women and girls. Among the high-density informal settlement residents of Lubaga Division of Kampala, where 78% of the population live in crowded single-room dwellings, informal daily wage labor, and with lack of personal financial independence, women are highly likely to be vulnerable to both household power dynamics and financial strife. In a recent community-based study of suburban Kampala communities, the overall prevalence of violence against women was 53.3% indicating that the population is at substantial risk of both multidimensional domestic gender-based violence (DGBV) and limitations in access to SRH services due to compounded factors such as intense economic hardship, reliance on fee-paying private clinics, and dependence on male partners [10–12].

However, despite the growing national acknowledgement of DGBV in Uganda, there remain notable evidence gaps at the micro-urban level. Existing evidence provides limited community-level understanding of the co-occurrence of physical, sexual, emotional and economic violence and their relationship with women’s sexual and reproductive health-seeking behaviour in informal urban settlements. There is also limited community based quantitative evidence to determine if specific types and cumulative burdens of DGBV can lead to certain SRH-seeking behaviours, namely condom usage, STI treatment-seeking, HIV testing and antenatal care. The following study has been designed to contribute to this gap by reporting: the prevalence of all types of DGBV; associated socio-demographic characteristics; and the relationship between exposure to each of these and all four types of SRH-seeking behaviour in a population of women living within the informal settlement of Lusaaze Zone, Lubaga Division, Kampala, Uganda.

## Methodology

### Study design

We conducted a quantitative, community-based analytical cross-sectional study rooted in a postpositivist framework of objective measurement and statistical inference. The study design involved the collection of data at one point in time to determine the prevalence and distribution of DGBV (descriptive study component). The study further attempts to examine the relationship between DGBV, sociodemographic correlates, and SRH-seeking behavior (analytical study component). Though this design is ideal for assessing burden of disease and violence, in this context where exposures and outcome are assessed at the same time, it was not able to establish causality.

### Study Area

This study was conducted in the Lusaaze Zone, Lubaga Division, Kampala District, Uganda. Lusaaze was chosen because of the suspected domestic and gender-based violence and reproductive health challenges in this population that could be driven by the economic pressures of kampala.

### Study population

The study population was women aged 15–49 years living in the Lusaaze Zone. The target population was estimated at 900–1,300 women from local administrative estimates and community records. The accessible population was women who had lived in Lusaaze for at least six months and who consented to take part (or assented with guardian consent, if aged under 18).

The sample population comprised all women in the age range of 15-49 years living in the Lusaaze Zone. An estimated target population of 900–1300 women was obtained from local administrative and the community registers. The participant population was women who had been residents in the study area for at least six months and who gave informed consent or assent (if aged under 18 years).

#### Inclusion criteria

Women aged 15–49 years, residence in the Lusaaze Zone for at least six months, informed consent, or assent with guardian consent for those under 18.

#### Exclusion criteria

Women who were critically ill at the time of data collection and women who were unable to communicate in English or Luganda during the interview.

### Sample size

The sample size for estimating a prevalence was calculated with the Kish and Leslie (1965) formula [13], n = Z²p(1 − p)/d², where Z = 1.96 (95% confidence), p = expected prevalence and d = 0.05 (margin of error). National data from the 2022 Uganda Demographic and Health Survey show that 43.7% of women have experienced physical violence since age 15 (Uganda Bureau of Statistics, 2023), while a local study in the suburbs of Kampala reported an overall violence prevalence of 53.3% [12]. The local figure was used:

n = (1.96)² × 0.533 × (1 − 0.533) / (0.05)² = 3.8416 × 0.2489 / 0.0025 = 382.5, rounded up to 383.

The calculated minimum sample size was 383 women. A total of 383 women aged 15–49 years were enrolled in the study.

### Sampling Method

Participants were recruited using a systematic random sampling technique. The sampling frame was compiled through a systematic enumeration of the residential households in the Lusaaze Zone with the assistance of local leaders and VHTs (Village Health Teams) resulting in 1,120 households identified as eligible for sampling from the Luzaase administrative register. The sampling interval, k, was calculated using the formula: k = N/n (1,120/383 = 2.92). The sampling interval was then rounded to three, k = 3. A starting household was randomly generated, k = 1-3 using a random number generator, and one household was surveyed and sampled along the geographic axis. The first household and subsequent homes were queried until reaching the sample size of 383 women.

Within each sampled household, one eligible woman between the ages of 15 and 49 was selected to participate. If more than one woman was eligible, participants were selected via a simple random ballot method with numbered slips of paper folded and placed in a bowl. If there was no eligible woman in a selected household, the next adjacent household was approached along the original geographic axis while not disturbing the original sampling frame. If a respondent was unavailable at the time of survey, up to two repeated visits were conducted at different points in the day before moving on to the next household.

### Data Collection

Using a structured instrument administered through interviews in either English or Luganda, as appropriate, data were obtained. The questionnaire was administered face-to-face and in a private room by trained research assistants, which was likely to minimize researcher bias and potentially improve the accuracy of responses for persons with low literacy levels and to allow for clarification of sensitive questions. The questionnaire covered three broad sections: (a) sociodemographic information, (b) domestic violence (i.e., physical, emotional, sexual, and economic violence) experiences and (c) sexual and reproductive health-seeking behaviors (i.e., antenatal care, family planning, HIV testing, and treatment-seeking for STI symptoms). Data were captured on mobile data-collection forms or printed questionnaires, with audio recordings not utilized.

Data were collected via a structured interview. The violence module asked specific questions about physical, sexual, emotional and economic violence. The questions on sexual, emotional and economic violence specifically asked about violence by the respondent’s current or most recent husband/partner and within the last 12 months. Questions on physical violence did not specifically ask about who the violence was from or when it occurred. Non-partner sexual violence was also asked about separately and was not included in the composite measure of DGBV.

### Validity and reliability

Content validity was established through a detailed review by academic supervisors and other public health research experts. Before the beginning of data collection, the pretested questionnaire in 20 women of a neighboring community with comparable sociodemographic settings, and the unclear issues were modified. Also, the interviewers got training on conducting interviews, ethical issues, and assuring privacy of the respondents which is most likely to reduce respondent and interviewer bias.

#### Exposure

Domestic gender based violence was measured using four categories: physical, sexual, emotional and economic violence. A positive answer to at least one question in each category indicated exposure to that type of violence. Physical and sexual violence were indicated by any answer other than ‘never’. Emotional and economic violence were indicated by positive answers. Sexual, emotional and economic violence questions asked about experiences with the respondents current or last husband/partner in the last 12 months. Physical violence questions did not ask about the perpetrators or time frame. Caution should be taken when interpreting this composite measure as there are differences in perpetrator and time frame between categories. Non-partner sexual violence was also measured.

Outcomes: SRH-Seeking behavior

Four sexual and reproductive health (SRH) main indicators and one exploratory indicator were analyzed. The indicators were: (1) whether the respondents have ever tested for HIV (yes/no); (2) whether they used antenatal care (ANC) during pregnancy (only for women who ever got pregnant), (3) whether they currently use any contraceptive method, compared to no method of use or abstain (yes/no); (4) whether the partner was a factor in family planning decision (has a partner ever kept them from using family planning?; only for women who ever had a partner), and (5) an exploratory indicator of women with at least one STI symptom seeking treatment for STI (for women with any symptoms, yes/no).

Covariates and Sociodemographic Variables. Respondent age group (15–24, 25–34, 35–49 years), highest education level (tertiary, secondary, none/primary), employment status (formally employed, self-employed, unpaid family helper, student), and partnership status (currently partnered, previously partnered, never partnered) were used to describe the sample and adjust multivariable models. Women who reported current or past partnerships were classified as “ever-partnered.”

### Data management and cleaning

Data were entered and validated at the time of collection; electronic data files were password protected, and paper questionnaires were stored in locked cabinets available only to the research team. The raw data file (n=383) was cleaned prior to analysis in R: (1) duplicate records were checked for and not identified, (2) spelling, capitalization and spacing were standardised, (3) resident eligibility was validated (women who had been in Lusaaze<6 months were excluded, as well as 3 records with missing length of residency, giving a final primary analytical sample of 377 women), (4) eight inconsistent response sets were recorded as missing (e.g., “agree” and “strongly disagree” responses to the same question) along with one implausible value, (5) the exposure, outcome and covariate variables were derived according to sections above, and (6) a single coded column was retained for each item and indicator variables were removed to prevent replication, as well as the “askid” identifier for each item. Missing data at item level was 2% across variables included in the analysis, and was dealt with using available-case analysis (model-by-model complete case analysis).

### Data Analysis

Statistical analyses were performed in R (version 4.3), with key outputs independently verified in Python. Descriptive statistics (frequencies and percentages) summarized respondent sociodemographic characteristics, forms of DGBV, and SRH-seeking indicators, with 95% confidence intervals (CIs) for proportions calculated using the Wilson score method.

For bivariate and multivariable analyses, modified Poisson regression with robust (sandwich) standard errors was used to estimate crude and adjusted prevalence ratios (cPR and aPR) with 95% CIs [14]. Modified Poisson regression was selected because all outcomes exceeded 10% prevalence, a setting in which logistic regression odds ratios substantially overestimate prevalence ratios. For Objective 2, each SRH-seeking indicator was modeled on DGBV exposure, adjusting for age group, education, employment status, and partnership status. Partner interference with family planning was analyzed specifically among ever-partnered women, while STI treatment-seeking was evaluated as an exploratory outcome restricted to women reporting at least one STI symptom. For Objective 3, any DGBV exposure was modeled simultaneously against age group, education, employment status, and partnership status. Additionally, a sensitivity analysis was conducted redefining the outcome as non-economic violence (physical, sexual, or emotional violence only) to evaluate its association with partner financial control without circularity. Statistical significance was set at a two-sided 0.05.

### Ethical Considerations

The study received ethics approval from the Uganda Heart Institute Research Ethics Committee (REC: UHIREC0192) with institutional clearance by Victoria University and administrative permission from the Lusaaze Zone Local Chairperson in Lubaga Division, Kampala. The researcher adhered to the ethical principles of the Declaration of Helsinki which include respect for persons, voluntary participation, anonymity and confidentiality, beneficence and justice. Written informed consent was obtained from adult participants before conducting interviews; written informed assent was obtained from 15–17 year olds in conjunction with written informed consent from their parents or guardians. Participants were appropriately informed of the purpose of the study, its risks and benefits, and that their participation was voluntary and they could refuse or withdraw at any time, without penalty. Interviews were conducted in private to ensure privacy and confidentiality, names were not recorded and participants were given identifiers on each record. Participants experiencing distress were treated with dignity and referred to local health and psychosocial services. Data was stored securely in password protected files and filing cabinets for use in the research and academic work.

## RESULTS

### Analytic sample and data completeness

The dataset contained 383 interview records. Three records reported residence in Lusaaze for less than six months and were excluded from Analysis. Residence duration was missing for a further 3 records; these were excluded from the analysis as well because eligibility could not be confirmed. The primary analytic dataset therefore comprised 377 women with confirmed residence of at least six months. Complete data across the four violence domains were available for 374 women.

### Sociodemographic characteristics of respondents

The largest age group was 35–49 years. Most respondents had a partner; tertiary education and formal employment were the most common categories (as shown in Table 1).

**Table 1.**
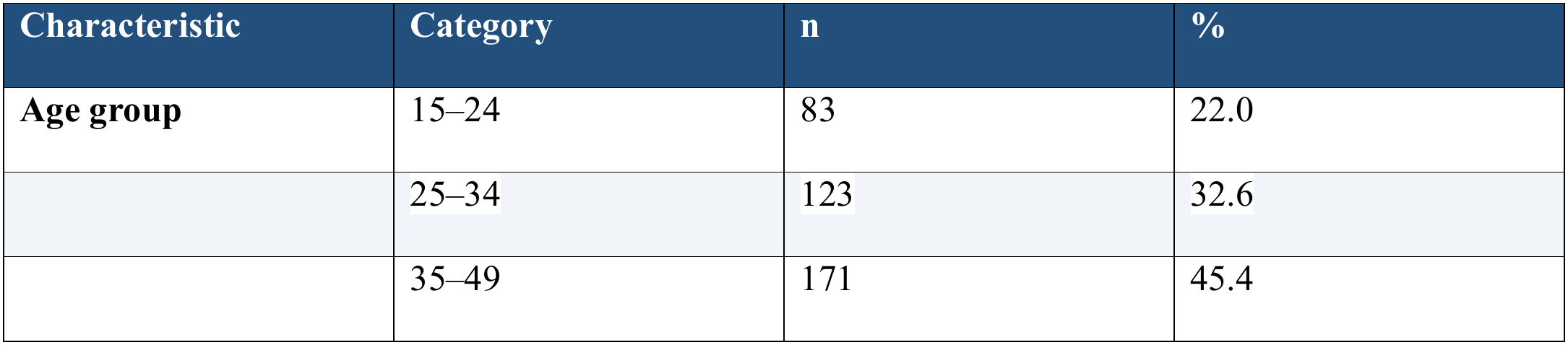
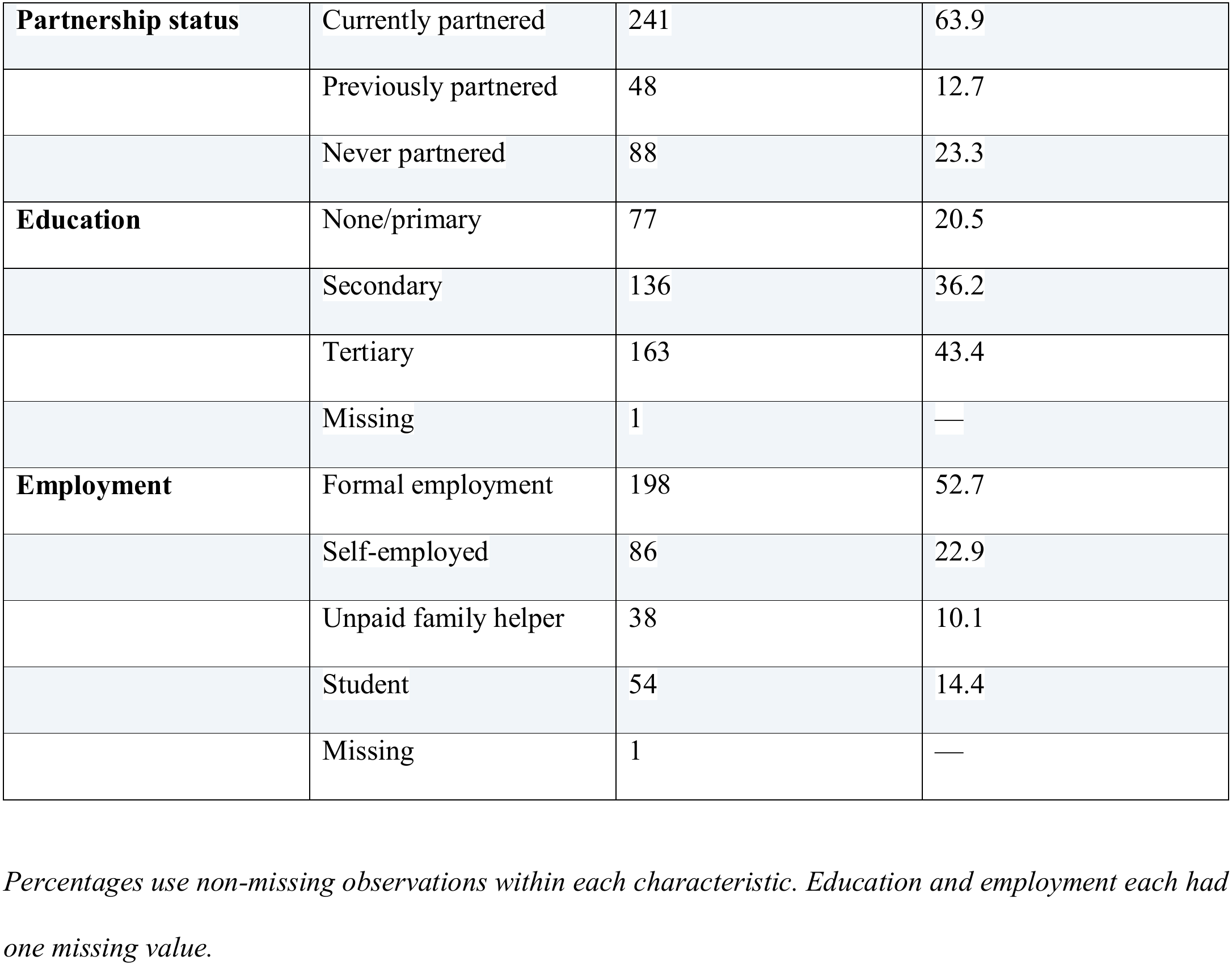
Sociodemographic characteristics of respondents (N = 377)

| Characteristic | Category | n | % |
|---|---|---|---|
| **Age group** | 15–24 | 83 | 22.0 |
|  | 25–34 | 123 | 32.6 |
|  | 35–49 | 171 | 45.4 |
| **Partnership status** | Currently partnered | 241 | 63.9 |
|  | Previously partnered | 48 | 12.7 |
|  | Never partnered | 88 | 23.3 |
| **Education** | None/primary | 77 | 20.5 |
|  | Secondary | 136 | 36.2 |
|  | Tertiary | 163 | 43.4 |
|  | Missing | 1 | — |
| **Employment** | Formal employment | 198 | 52.7 |
|  | Self-employed | 86 | 22.9 |
|  | Unpaid family helper | 38 | 10.1 |
|  | Student | 54 | 14.4 |
|  | Missing | 1 | — |

*Percentages use non-missing observations within each characteristic. Education and employment each had one missing value.*

### Prevalence and forms of domestic gender-based violence

Among 374 women with complete data across the four violence domains, 271 (72.5%) reported at least one form of violence (95% CI 67.7–76.7%). This composite reflects the questionnaire’s four-domain classification and should not be interpreted as a uniform past-year estimate of intimate partner violence. Emotional violence was the most frequently reported form, followed by physical, sexual and economic violence (see table 2 and Figure 1).

**Figure 1.**
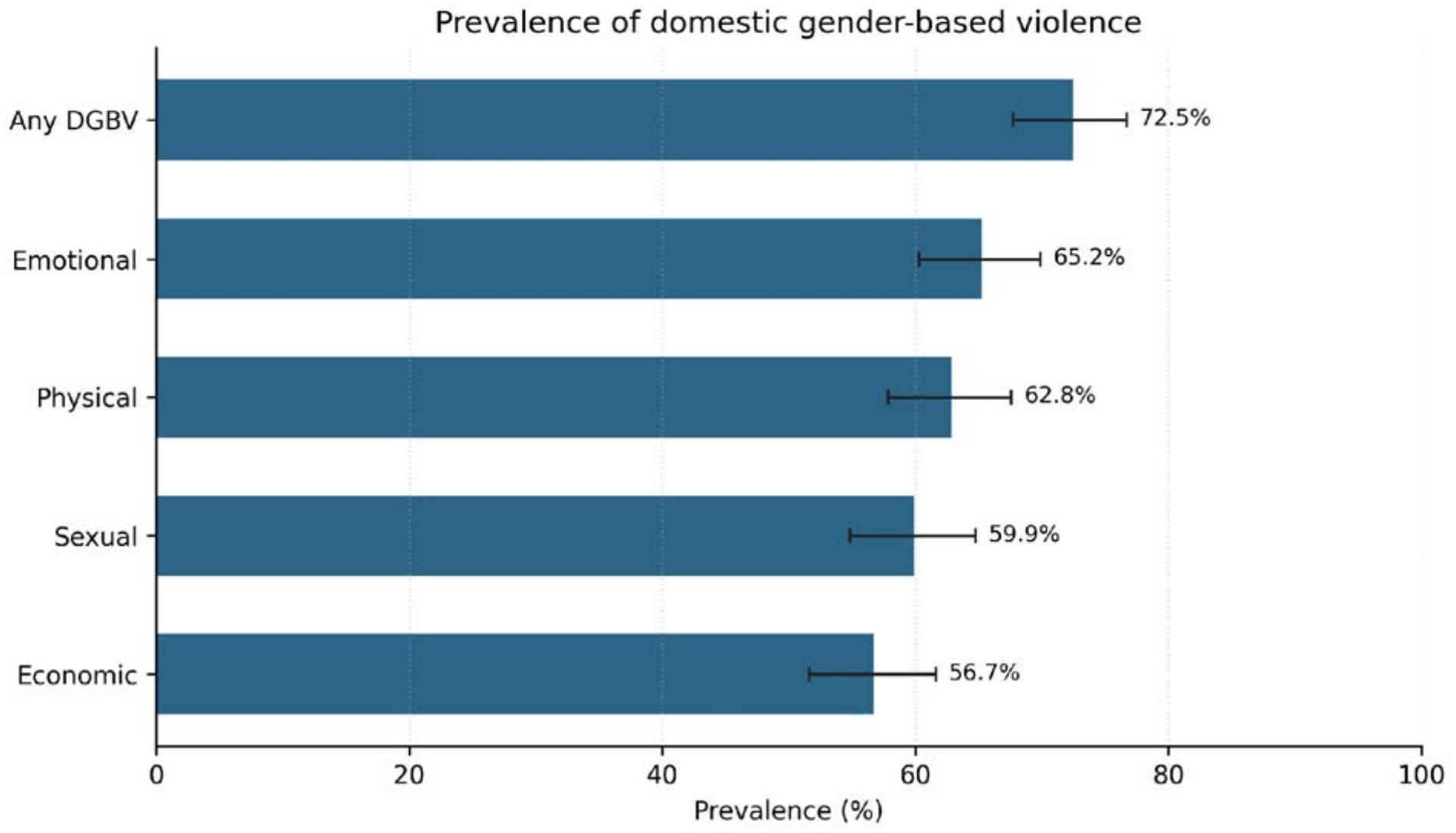
Prevalence of domestic gender-based violence by form with 95% Wilson confidence intervals. Of the 374 women with complete data across all four forms, 103 (27.5%) reported none, 30 (8.0%) one form, 24 (6.4%) two forms, 31 (8.3%) three forms and 186 (49.7%) all four forms (see table 3)

**Table 2.** Prevalence of domestic gender-based violence by form.

| **Indicator** | **n/N** | **Prevalence** | **95% CI** |
|---|---|---|---|
| Any domestic gender-based violence | 271/374 | 72.5% | 67.7–76.7% |
| Physical violence | 235/374 | 62.8% | 57.8–67.6% |
| Sexual violence | 224/374 | 59.9% | 54.8–64.7% |
| Emotional violence | 244/374 | 65.2% | 60.3–69.9% |
| Economic violence | 212/374 | 56.7% | 51.6–61.6% |

*Confidence intervals are 95% Wilson intervals. A domain was positive if at least one item was endorsed; physical and sexual items were positive for responses other than 'never', and emotional/economic items for 'yes'.*

**Table 3.**
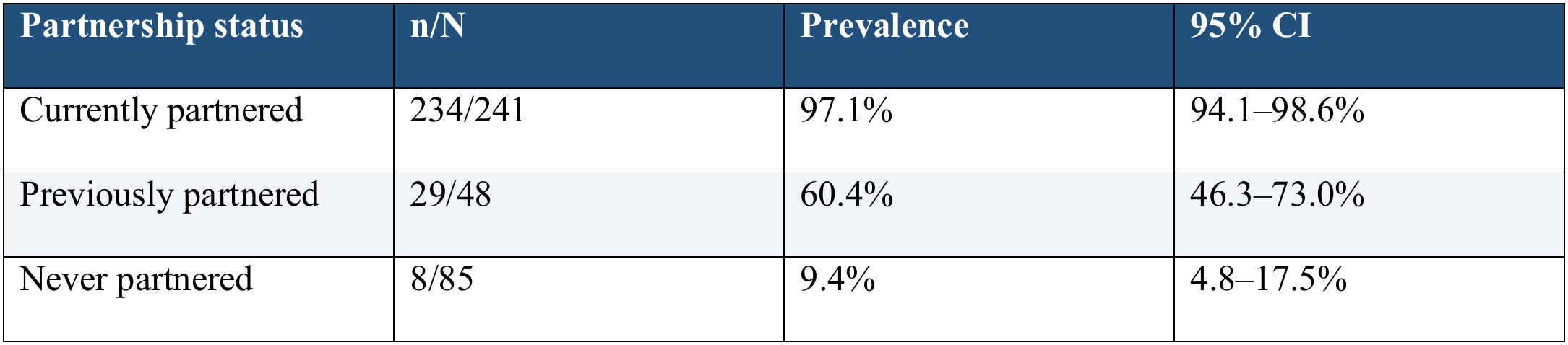
Prevalence of any domestic gender-based violence by partnership status.

**Table 3.1:** Acts used to define each form of domestic gender-based violence.

| **Form** | **Number of acts** | **Acts included** |
|---|---|---|
| Physical | 6 | Pushed, shaken or had something thrown at her; slapped or arm twisted; punched; kicked, dragged or beaten; choked or burned; threatened with a weapon |
| Sexual | 3 | Forced intercourse; forced other sexual acts; threatened to force sex |
| Emotional | 7 | Humiliated in public; threatened with harm; insulted; intimidated; threatened about children or support; accused of unfaithfulness; contact with others restricted |
| Economic | 5 | Refused money; earnings taken; forced to work; prevented from working; partner made financial decisions alone |

### Association between DGBV and SRH-seeking indicators

SRH indicators were analyzed using outcome-specific eligible populations. Modified Poisson regression with robust (sandwich) standard errors was used to estimate prevalence ratios. Adjusted models included age group, education, employment and partnership status. For partner interference with family planning, analysis was restricted to ever-partnered women with a valid yes/no response. STI symptom/treatment findings are exploratory because symptom endorsement was unusually high (see table 4).

**Table 4.** SRH-seeking indicators by DGBV exposure and modified Poisson regression estimates.

| **Outcome** | **Overall n/N (%)** | **DGBV exposed n/N (%)** | **DGBV unexposed n/N (%)** | **Crude PR (95% CI)** | **Crude p** | **Adjusted PR (95% CI)** | **Adjusted p** |
|---|---|---|---|---|---|---|---|
| STI treatment among women reporting ≥1 symptom (exploratory) | 335/356 (94.1%) | 248/264 (93.9%) | 84/89 (94.4%) | 1.00 (0.94–1.06) | 0.876 | 1.00 (0.90–1.12) | 0.937 |
| Ever tested for HIV | 316/375 (84.3%) | 237/269 (88.1%) | 76/103 (73.8%) | 1.19 (1.06–1.35) | 0.005 | 0.99 (0.82–1.20) | 0.938 |
| ANC attendance among ever-pregnant women | 305/310 (98.4%) | 259/262 (98.9%) | 44/46 (95.7%) | 1.03 (0.97–1.10) | 0.305 | 0.93 (0.84–1.02) | 0.132 |
| Current contraceptive use | 363/377 (96.3%) | 268/271 (98.9%) | 92/103 (89.3%) | 1.11 (1.03–1.19) | 0.003 | 1.11 (1.01–1.22) | 0.029 |
| Partner prevented family planning among ever-partnered women | 134/278 (48.2%) | 131/260 (50.4%) | 3/18 (16.7%) | 3.02 (1.07–8.55) | 0.037 | 3.36 (1.25–9.06) | 0.017 |

*PR = prevalence ratio; CI = confidence interval. Adjusted models included age group, education, employment and partnership status. Overall denominators are outcome-specific; exposed/unexposed estimates use records with known DGBV status. STI treatment is exploratory.*

After adjustment, DGBV was not clearly associated with HIV testing (aPR 0.99, 95% CI: 0.82 – 1.20) or ANC attendance among ever-pregnant women (aPR 0.93, 95% CI: 0.84 – 1.02). Current contracepti e use was slightly more prevalent among women reporting DGBV (aPR 1.11, 95% CI: 1.01–1.22), although this estimate should be interpreted cautiously because only 14 women reported no current method or abstinence. Among ever-partnered women with a valid response, partner interference with family planning was reported by 50.4% of women classified as exposed to DGBV and 16.7% of those not classified as exposed; the adjusted prevalence ratio was 3.36 (95% CI: 1.25–9.06). This estimate was imprecise because only 18 unexposed ever-partnered women contributed to the comparison (see figure 2). In the exploratory STI analysis, 335/356 (94.1%) of symptomatic women reported treatment-seeking. However, 356 of 377 women (94.4%) reported at least one of the four listed symptoms, and individual symptom endorsement was also high. Because the reference period and clinical meaning of these symptom items are unclear, this result should not be interpreted as a reliable estimate of STI morbidity or definitive evidence about STI- care seeking.

**Figure 2.**
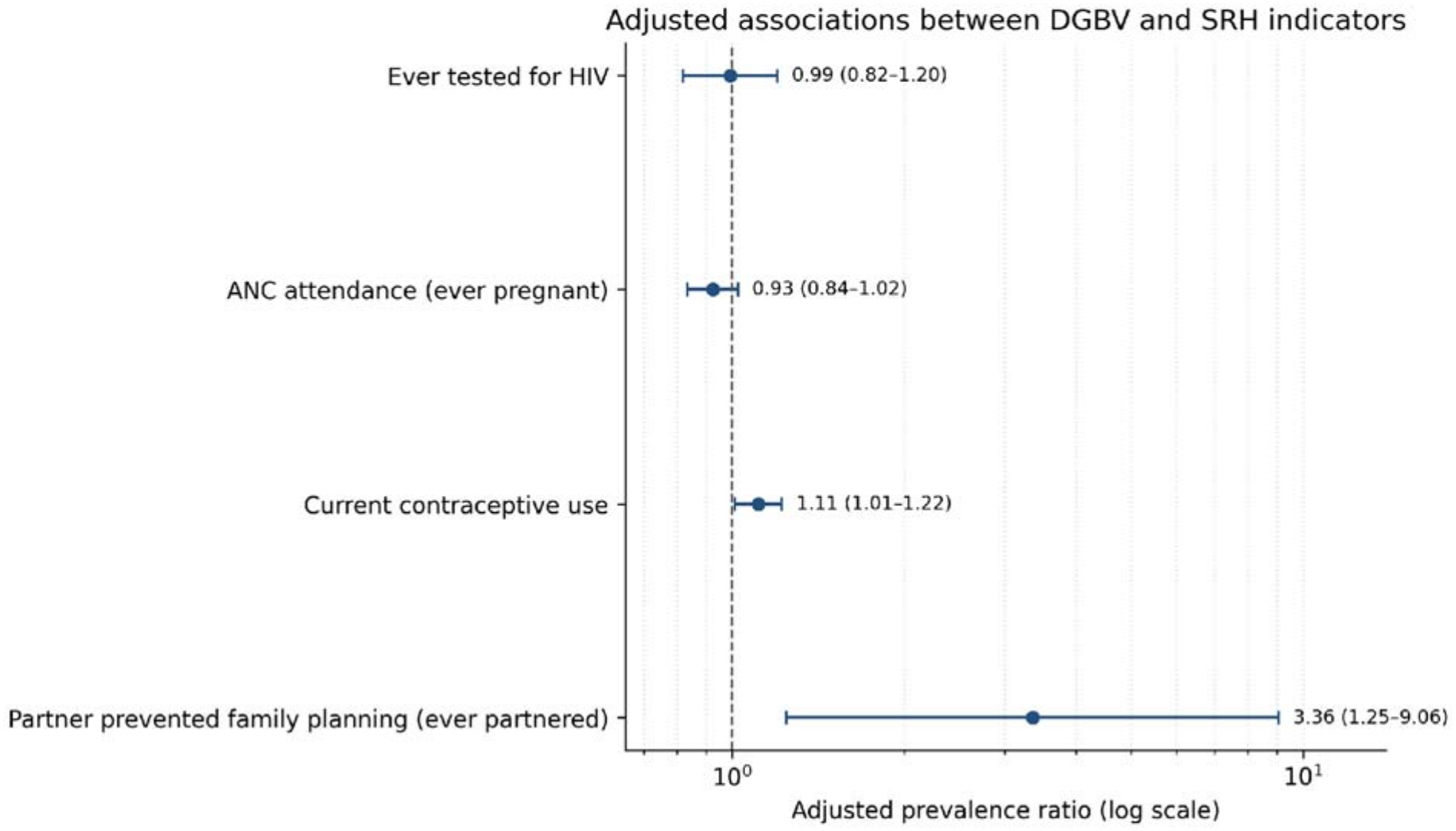
Adjusted prevalence ratios for four principal SRH indicators. Dashed vertical line marks PR = 1; STI treatment is omitted from the figure because of measurement concerns.

### Factors associated with DGBV

Modified Poisson regression was used to estimate crude and adjusted prevalence ratios for any DGBV. The adjusted model included age group, education, employment and partnership status. Partnership status showed the strongest association with DGBV classification. After adjustment, age was not clearly associated with DGBV; lower education and self-employment showed modest positive associations, while the student estimate was imprecise (see Table 5).

**Table 5.** Crude and adjusted factors associated with any domestic gender-based violence.

| **Factor** | **Category** | **Exposed n/N (%)** | **Crude PR (95% CI)** | **Crude p** | **Adjusted PR (95% CI)** | **Adjusted p** |
|---|---|---|---|---|---|---|
| Age group | 25–34 | 97/123 (78.9%) | 2.87 (1.99–4.14) | <0.001 | 0.96 (0.83–1.11) | 0.603 |
| Age group | 35–49 | 152/171 (88.9%) | 3.23 (2.26–4.63) | <0.001 | 1.00 (0.86–1.17) | 0.971 |
| Education | Secondary | 101/134 (75.4%) | 1.08 (0.94–1.24) | 0.279 | 1.12 (1.04–1.21) | 0.004 |
| Education | None/primary | 56/77 (72.7%) | 1.04 (0.88–1.24) | 0.631 | 1.13 (1.02–1.26) | 0.020 |
| Employment | Self-employed | 78/86 (90.7%) | 1.22 (1.10–1.36) | <0.001 | 1.08 (1.00–1.16) | 0.048 |
| Employment | Unpaid family helper | 35/38 (92.1%) | 1.24 (1.10–1.40) | <0.001 | 0.99 (0.90–1.08) | 0.804 |
| Employment | Student | 10/51 (19.6%) | 0.26 (0.15–0.46) | <0.001 | 1.74 (0.95–3.21) | 0.074 |
| Partnership status | Previously partnered | 29/48 (60.4%) | 0.62 (0.49–0.78) | <0.001 | 0.58 (0.45–0.73) | <0.001 |
| Partnership status | Never partnered | 8/85 (9.4%) | 0.10 (0.05–0.19) | <0.001 | 0.07 (0.04–0.13) | <0.001 |

*Reference categories: age 15–24 years; tertiary education; formal employment; currently partnered.*
*Adjusted models included all four covariates. Estimates are associations, not causal effects.*

In the adjusted model (n = 372), women with secondary education had a higher prevalence of DGBV than women with tertiary education (aPR 1.12, 95% CI 1.04–1.21), as did women with no formal/primary education (aPR 1.13, 95% CI 1.02–1.26). Compared with currently partnered women, prevalence was lower among previously partnered women (aPR 0.58, 95% CI: 0.45 – 0.73) and never-partnered women (aPR 0.07, 95% CI: 0.04 – 0.13). These associations should be interpreted in light of the relationship between partnership history and opportunity for partner violence, and the small number of unexposed currently partnered women (see table 5).

In a sensitivity analysis using physical, sexual and emotional violence only (excluding all economic-violence items), 270/376 women (71.8%) met the definition. Financial control was associated with this non-economic violence outcome (aPR 1.14, 95% CI 1.06 – 1.23; p <0.001) after adjustment for age, education, employment and partnership status. This sensitivity analysis avoids using the financial-control item as part of the outcome it predicts.

### Summary of findings

Among 374 women with complete data across the four violence domains, 271 (72.5%) reported at least one form of violence. This composite reflects the questionnaire’s four-domain classification and should not be interpreted as a uniform past-year estimate of intimate partner violence and 49.7% reported all four forms. After adjustment, DGBV was not clearly associated with HIV testing or ANC attendance; current contraceptive use was modestly higher, and partner interference with family planning was more frequently reported among exposed ever-partnered women. Partnership status showed the strongest association with the composite violence classification; however, this finding should be interpreted cautiously because three violence domains were measured specifically in relation to current or former partners. STI symptom/treatment findings remain exploratory because of unusually high symptom endorsement and unclear reference period.

## Discussion

This study identified a high prevalence of reported violence across four domains among women living in Lusaaze, Kampala. However, differences in perpetrator attribution and recall periods across the questionnaire domains require caution when interpreting the composite estimate as domestic or intimate partner violence. In general (72.5%) of the surveyed women who had completed the full battery of questions in each of the four DGBV questionnaires reported experiencing at least one form of DGBV. The percentage of women reporting experience of DGBV was significantly higher than previous estimates from national surveys conducted in Uganda, such as Uganda’s Demographic and Health Surveys, which estimated that thirty-five to fifty percent of ever married women experienced either physical or sexual violence during the year preceding the survey [15, 16]. Nonetheless, these national survey estimates cannot be compared directly to those found in this study due to many methodological differences. For example, national surveys employ stratified random samples from a large geographic area —commonly including rural areas while this study used systematic sampling of a small geographic area, a single peri-urban informal settlement.

Additionally, national surveys typically include items assessing physical and sexual violence only over a specific time period —e.g., past twelve month while this study included behavior-specific items assessing physical, sexual, emotional and economic violence. The inclusion of behavioral items assessing emotional and economic violence may have contributed to increasing the number of cases detected in this study relative to studies that assess only physical and sexual violence. Evidence exists suggesting that assessment tools focusing on both physical and sexual forms of violence underreport experiences of other types of violence when compared to assessment tools designed to measure all forms of violence [18, 19]. A further contributing factor to the high levels of reported DGBV in informal communities could be the presence of socioeconomic disadvantages and limited access to potential sources of support services. While the current study did not collect sufficient data to determine whether these factors were causal determinants of the high rates of DGBV reported by women in this study [20, 21], they are likely to have been relevant factors.

Emotional abuse was the most frequent type of violence reported at 65.2%. The second highest frequency was for physical abuse at 62.8%, followed by sexual abuse at 59.9% and then economic abuse at 56.7%. These rates are similar to other reports from sub-Saharan Africa and South Asia that report psychological abuse, either alone or with more than physical abuse. In addition, nearly half (49.7%) of the sample were exposed to all four types of violence, suggesting that women in this sample were likely to experience a pattern of multiple exposures of violence.

When victims of violence experience two or more types of violence simultaneously it can be referred to as polyvictimization. Polyvictimization is a serious issue since experiencing different forms of victimization can have additive negative impacts on mental and physical health [17, 24]. Of note, there were significantly greater percentages of currently partnered women (97.1%) who reported having experienced one or more forms of DGBV compared to previously partnered women (60.4%) and never partnered women (9.4%). The higher prevalence of reported violence among currently partnered women highlights the importance of examining relationship context, while recognising that the study did not establish whether the current partner was responsible for every reported physical violence experience [25, 26]. Although the rates of abuse among never partnered women are high, they should be interpreted cautiously. There could be several explanations for these high rates, such as being abused by someone else in the same household or being abused in a previous relationship. However, whether these abuses occurred in previous relationships will depend upon how the investigators define and measure abuse in the DGVB survey. Abuse can occur against a woman by many people outside her intimate partner(s) whereas intimate partner violence refers only to violence from an individual’s present or past intimate partner(s) [27, 28]. Understanding this difference is key to comparing our results with studies examining only partner specific violence, such as the Kenya DHS examination of intimate partner based sexual violence and the South African examination of physical violence by present or past partner(s) [25, 29].

There was not a consistent relationship found between DGBV and utilization of SRH services. As indicated earlier, we did find a statistically non-significant association between DGBV and use of HIV testing (aPR 0.99, 95% CI: 0.82 – 1.20). Our finding contradicts those from an intervention trial in Nairobi, Kenya where screening for intimate partner violence during HIV counseling resulted in increased self-reporting of both HIV testing and use of HIV related services [30]. However, the studies differ substantially in design and setting: the Nairobi study evaluated an intervention among clinic attendees, whereas the present study examined community-level associations. Furthermore, HIV testing coverage in Lusaaze was already high (84.3%), potentially limiting the observable variation needed to identify differences between exposure groups. Violence may also influence testing in opposing directions: fear of partner retaliation may discourage testing, while perceived HIV risk or the need for reproductive health information may motivate it [31–33]. Consequently, the absence of an association should not be interpreted as evidence that violence has no influence on HIV testing decisions.

DGBV was not significantly associated with ANC attendance (aPR 0.93, 95% CI: 0.84-1.02). This contrasts with evidence from systematic reviews and meta-analyses that intimate partner violence is associated with reduced adequate utilisation of ANC services [34,35]. However, the near universal attendance of every pregnant woman (98.4%) may explain why this association was not significant due to limited variation. Evidence from a 36 country analysis of DHS data suggests that lifetime intimate partner violence is more consistently associated with reduced completion of recommended ANC visits and delivery at a facility [36]. However, the measure of whether women attended ANC at least once may not capture the full picture of how intimate partner violence impacts on women’s ANC. In high initial contact settings, intimate partner violence may impact on women’s ability to adequately access and utilise services [36]. Therefore, the high ANC attendance in Lusaaze should not be taken to mean that women are accessing equitable and violence free maternal health services.

A notable finding was the modest positive association between DGBV and current contraceptive use (aPR 1.11, 95% CI: 1.01-1.22). This is contrary to findings showing that women exposed to intimate partner violence have reduced contraceptive use [23, 37]. However, this is consistent with findings from Bawuah et al (38) who found that women experiencing intimate partner violence had higher modern contraceptive use but lower adequate maternal health utilisation from a 25 country DHS analysis in Africa. This suggests that contraceptive use and maternal health utilisation may be impacted in opposing ways by relationship control. One possible explanation for this is that women are using contraceptives discreetly. Studies from Uganda and Bangladesh have shown that women are using discreet contraceptive methods to retain some control over their reproduction within controlling or abusive relationships [39,40]. This could therefore explain the association seen in Lusaaze. Another factor to consider is whether the association is due to reverse causation. Women using contraception may cause suspicion from their partner. Longitudinal and qualitative studies are needed to determine if women are making autonomous decisions around contraceptive use or if they are using discreet contraceptive methods. It should be noted that only 14 women were non-users so this may have impacted on the stability of the results [44].

The strongest association with an SRH outcome was partner interference with family planning (aPR 3.36, 95% CI: 1.25-9.06). Nearly half of ever partnered women (48.2%) experienced interference with family planning. Women who experienced DGBV had over 3 times higher prevalence of partner interference. This suggests that reproductive coercion is an important aspect of violence experienced in Lusaaze. The reproductive coercion framework explains how women experience sabotage of contraceptive use, pressure to conceive and interference with contraceptive decision making from their partner which limits their reproductive autonomy [41, 45]. This is consistent with qualitative findings from Wakiso District in Uganda where women reported that their partners would break contraceptive pills, refuse to use condoms and pressure them to conceive [39]. Similar findings have been seen in Bangladesh and India where women have reported partner and family resistance including in-laws to contraceptive use [40, 37]. However, the wide confidence interval suggests that there is a lot of uncertainty around the strength of this association. There were only a small number of ever partnered women who were not exposed (n=18). This is an issue with sparse data and can be particularly problematic in areas where prevalence is high. This reduces the stability of the adjusted estimates [46,47].

The sociodemographic findings show that DGBV is influenced by a range of individual and relationship factors. Women with no or primary education (aPR 1.13) and secondary education (aPR 1.12) had higher prevalence of DGBV compared to those with tertiary education. This is generally consistent with evidence showing women with lower levels of education are more vulnerable to intimate partner violence [48, 49]. However, there was not a large difference so it cannot be said that women with tertiary education are protected from DGBV. Education may be an indicator of wider socioeconomic differences, economic opportunities and power in relationships. It may also influence whether women are able to identify, report and leave abusive relationships [50, 51]. Women who were self-employed had higher prevalence of DGBV (aPR 1.08). Whilst this may increase women’s financial independence it does not necessarily increase their power in relationships. Women may challenge traditional gender norms by becoming economically active which can cause conflict and violence [52,53]. This shows the importance of understanding violence beyond just individual socioeconomic factors and considering how gendered power and resources are distributed [54,55].

Age was not significantly associated with DGBV. This is contrary to studies showing that younger women are more likely to experience violence [56,57]. However, this is not necessarily conflicting as there is evidence that women experience violence throughout their reproductive life. This may also be due to the high proportion of women aged between 35-49 years old (45.4%). There may also be differences in relationship transitions such as separation and widowhood.

The findings regarding STI symptoms should be treated with caution. The high prevalence of symptoms (94.4%) and seeking treatment (94.1%) may not be accurate due to issues with how symptoms were categorised. Symptoms such as discharge and itch are not specific to STIs and could be caused by other gynaecological or physiological issues. This has been seen in other studies using symptoms as a method of screening [59,60]. Therefore, it cannot be confidently said that there is no association between DGBV and seeking treatment for STIs as the outcome may not accurately reflect true STI morbidity or appropriate treatment. Future studies should include lab confirmed STI testing or validated syndromic algorithms with clear time frames [18].

Overall, the findings can be best understood using the social ecological model, feminist perspectives and reproductive coercion. The social ecological model considers that violence occurs due to a combination of individual, relationship and community factors. Feminist perspectives consider power imbalances in relationships to control women’s bodies and reproduction [55, 54, 61]. Reproductive coercion explains how women’s reproduction is controlled by their partner through interfering with contraceptive use and conception decisions [41, 45]. The high prevalence of contraceptive use combined with high partner interference shows that traditional measures of service use may not fully reflect women’s reproductive autonomy. Women may be accessing contraception but still experience limited control over their reproduction. This demonstrates that women may not be empowered, making informed decisions or free from coercion [39, 40, 62].

This should be interpreted with consideration to the limitations. The study was cross-sectional meaning that it is not possible to determine causation or temporality. There was also no clear time frame for violence meaning that the results may not be as accurate. Non-response may have caused bias. There were issues with the validity of the STI questions and the exact measure of violence. There were a limited number of currently partnered women who were not exposed meaning some of the adjusted estimates were not as accurate. Particularly the association between partner interference with family planning. However, the study provides evidence from a community setting in an under-researched informal urban area. It looked at a range of different types of violence and included specific behaviours. The decisions made during analysis were reported meaning the findings are transparent. From a public health perspective, interventions should go beyond increasing women’s service use and consider improving safety, confidentiality, reproductive autonomy and identifying partner interference. Women may not be empowered as they have high ANC attendance and contraceptive uptake but relationship coercion is still prevalent [36,63].

There were different ways of asking about perpetrators of violence. The questions on sexual, emotional and economic violence specifically asked about current or ex-husbands/partners in the last 12 months. Questions on physical violence were not specific about perpetrators or time frame. This means the 4 domain composite combines different time frames and cannot be viewed as a single measure of intimate partner violence in the last year. Future studies should be longitudinal and include validated reproductive coercion measures. Lab confirmed STI testing should also be included. The quality, continuity and autonomy of care should also be considered. This would provide better evidence to implement integrated gender sensitive interventions to improve women’s SRH in informal urban settlements.

## Conclusion

Violence exposure across multiple domains was common among women in Lusaaze. The coexistence of contraceptive use and partner interference with family planning highlights the limitations of healthcare utilisation as a standalone indicator of reproductive autonomy. Survivor-centred SRH services should address violence, confidentiality and reproductive decision-making.

### Recommendations

SRH services should offer confidential, survivor focussed strategies to identify violence and reproductive coercion in informal urban settings. There should be clear referral routes for psychosocial, legal and protection services. Confidentiality should be maintained whilst supporting women to make their own decisions on contraception. Public health strategies should also be implemented to reduce violence in the community. Longitudinal and mixed methods studies should be conducted to understand the causal pathways. Validated tools should be used to measure violence and reproductive coercion.

## Declarations

### Ethics approval

The study received ethics approval from the Uganda Heart Institute Research Ethics Committee (REC: UHIREC0192) with institutional clearance by Victoria University and administrative permission from the Lusaaze Zone Local Chairperson in Lubaga Division, Kampala. The researcher adhered to the ethical principles of the Declaration of Helsinki which include respect for persons, voluntary participation, anonymity and confidentiality, beneficence and justice. Written informed consent was obtained from adult participants before conducting interviews; written informed assent was obtained from 15–17 year olds in conjunction with written informed consent from their parents or guardians. Participants were appropriately informed of the purpose of the study, its risks and benefits, and that their participation was voluntary and they could refuse or withdraw at any time, without penalty. Interviews were conducted in private to ensure privacy and confidentiality, names were not recorded and participants were given identifiers on each record. Participants experiencing distress were treated with dignity and referred to local health and psychosocial services. Data was stored securely in password protected files and filing cabinets for use in the research and academic work.

## Data Availability

The datasets generated and analysed during the current study are not publicly available because of the sensitive nature of clinical and personal data relating to sexual, gender and reproductive health. Data is available from the corresponding author upon reasonable request, based on attached ethical and institutional conditions.

## Consent for publication

Not applicable. No individual person’s identifying data, images or videos are included in this manuscript. All participant data have been fully anonymised.

## Authors’ contributions

KS: Edited and wrote this paper, analyzed and submitted the final copy

NS: conceptualized and wrote the proposal of this study

## Acknowledgements

The authors sincerely compliment Victoria University and the administrative local councils of Lusaaze community in Rubaga division.

## References

1 World Health Organization: WHO. Violence against women. World Health Organization: WHO. 2026; published online June 18. https://www.who.int/news-room/fact-sheets/detail/violence-against-women (accessed Nov 12, 2026).

2 Heise LL. Violence Against Women: An Integrated, Ecological Framework. Violence Against Women 1998; 4: 262–90.

3 Postmus JL, Hoge GL, Breckenridge J, Sharp-Jeffs N, Chung D. Economic Abuse as an Invisible Form of Domestic Violence: A Multicountry Review. *Trauma, Violence*, & Abuse 2018; 21: 261–83.

4 World Health Organization. Prevalence of intimate partner violence and non-partner sexual violence against adolescent girls aged 15–19 years: technical brief. World Health Organization, 2026 10.2471/b09912 (accessed Nov 12, 2026).

5 Campbell JC. Health consequences of intimate partner violence. The Lancet 2002; 359: 1331–6.

6 Maxwell L, Devries K, Zionts D, Alhusen JL, Campbell J. Estimating the Effect of Intimate Partner Violence on Women’s Use of Contraception: A Systematic Review and Meta-Analysis. PLOS ONE 2015; 10: e0118234.

7 Muluneh MD, Stulz V, Francis L, Agho K. Gender Based Violence against Women in Sub-Saharan Africa: A Systematic Review and Meta-Analysis of Cross-Sectional Studies. International Journal of Environmental Research and Public Health 2020; 17: 903.

8 Nabaggala MS, Reddy T, Manda S. Effects of rural–urban residence and education on intimate partner violence among women in Sub-Saharan Africa: a meta-analysis of health survey data. BMC Women’s Health 2021; 21. DOI:10.1186/s12905-021-01286-5.

9 ubosadmin. Uganda Demographic and Health Survey 2022 Main Report. Uganda Bureau of Statistics. 2024; published online June 20. https://www.ubos.org/2024/06/20/uganda-demographic-and-health-survey-2022-main-report/ (accessed Nov 12, 2026).

10 ubosadmin. Gender Based Violence Thematic Report Based on UDHS 2022. Uganda Bureau of Statistics. 2026; published online March 6. https://www.ubos.org/2026/03/06/gender-based-violence-thematic-report-based-on-udhs-2022/ (accessed Nov 12, 2026).

11 Nakakawa J, Nanyingi M, Andinda M, Lwenge M. Prevalence and factors associated with health facility-based deliveries among urban slum settlers in Lubaga Division, Kampala District, Uganda: a community-based cross-sectional study. BMC Health Services Research 2026; 26. DOI:10.1186/s12913-026-14445-x.

12 Nalubuuka A, Ekung E, Nekesa C, Atim PG, Udho S. Association between intimate partner violence and utilization of antenatal care services among women in suburbs of Kampala, Uganda. Discover Public Health 2026; 23. DOI:10.1186/s12982-026-02037-3.

13 Survey Sampling. By Leslie Kish. (New York: John Wiley & Sons, Inc., 1965. Pp. xvi, 643. $10.95.). American Political Science Review 1965; 59: 1025–1025.

14 Zou G. A Modified Poisson Regression Approach to Prospective Studies with Binary Data. American Journal of Epidemiology 2004; 159: 702–6.

15 Ma N, Chen S, Kong Y, et al. Prevalence and changes of intimate partner violence against women aged 15 to 49 years in 53 low-income and middle-income countries from 2000 to 2021: a secondary analysis of population-based surveys. The Lancet Global Health 2023; 11: e1863–73.

16 Uganda Demographic and Health Survey 2016. TB DIAH. https://www.tbdiah.org/resources/publications/uganda-demographic-and-health-survey-2016/ (accessed Nov 12, 2026).

17 Potter LC, Morris R, Hegarty K, García-Moreno C, Feder G. Categories and health impacts of intimate partner violence in the World Health Organization multi-country study on women’s health and domestic violence. International Journal of Epidemiology 2020; 50: 652–62.

18 Meyer SR, Stöckl H, Vorfeld C, Kamenov K, García-Moreno C. A scoping review of measurement of violence against women and disability. PLOS ONE 2022; 17: e0263020.

19 Stöckl H, Penhale B. Intimate Partner Violence and Its Association With Physical and Mental Health Symptoms Among Older Women in Germany. Journal of Interpersonal Violence 2014; 30: 3089– 111.

20 Kovacs RJ. The macro-level drivers of intimate partner violence: New evidence from a multilevel dataset. Global Public Health 2017; 13: 944–56.

21 Messner W. Beyond the individual: global socioeconomic, cultural, and religious contexts of male-perpetrated intimate partner violence. World Development 2025; 195: 107121.

22 Dangal G, Shrestha N, Khanal G, et al. Prevalence and Contributing Factors of Gender-based Violence in SAARC Territories from 2010 to 2020: A Systematic Review and Meta-analysis. Journal of Nepal Health Research Council 2022; 20: 1–11.

23 Shrestha SK, Thapa S, Vicendese D, Erbas B. Women’s attitude towards intimate partner violence and utilization of contraceptive methods and maternal health care services: an analysis of nationally representative cross-sectional surveys from four South Asian countries. BMC Women’s Health 2022; 22. DOI:10.1186/s12905-022-01780-4.

24 Ellsberg M, Heise L. Bearing witness: ethics in domestic violence research. The Lancet 2002; 359: 1599–604.

25 Jewkes R, Levin J, Penn-Kekana L. Risk factors for domestic violence: findings from a South African cross-sectional study. Social Science & Medicine 2002; 55: 1603–17.

26 Melkam M, Fentahun S, Rtbey G, et al. Multilevel analysis of intimate partner violence and associated factors among reproductive-age women: Kenya demographic and health survey 2022 data. BMC Public Health 2024; 24. DOI:10.1186/s12889-024-19012-9.

27 Morrison A, Ellsberg M, Bott S. Addressing Gender-Based Violence: A Critical Review of Interventions. The World Bank Research Observer 2007; 22: 25–51.

28 Jordan CE. DOMESTIC VIOLENCE. In: Encyclopedia of Forensic and Legal Medicine. Elsevier, 2005: 223–9.

29 Ikwara EA, Humphrey A, Meble K, Hannington WG, Jacob EC, Isiko I. Prevalence and factors influencing intimate partner sexual violence against women aged 15–49 in Kenya: findings from the 2022 Kenya demographic and health survey. BMC Women’s Health 2025; 25. DOI:10.1186/s12905-025-03593-7.

30 Haberland N, Ndwiga C, McCarthy K, et al. Addressing Intimate Partner Violence and Power in Intimate Relationships in HIV Testing Services in Nairobi, Kenya. AIDS and Behavior 2020; 24: 2409–20.

31 Leight J, Wilson N. Intimate partner violence and maternal health services utilization: evidence from 36 National Household Surveys. BMC Public Health 2021; 21. DOI:10.1186/s12889-021-10447-y.

32 Chibber KS, Krishnan S. Confronting Intimate Partner Violence: A Global Health Priority. Mount Sinai Journal of Medicine: A Journal of Translational and Personalized Medicine 2011; 78: 449–57.

33 Parsons L, Goodwin MM, Petersen R. Violence Against Women and Reproductive Health: Toward Defining a Role for Reproductive Health Care Services. Maternal and Child Health Journal 2000; 4: 135– 40.

34 Musa A, Chojenta C, Geleto A, Loxton D. The associations between intimate partner violence and maternal health care service utilization: a systematic review and meta-analysis. BMC Women’s Health 2019; 19. DOI:10.1186/s12905-019-0735-0.

35 Liu L, Liang D, Anwar S, et al. Overlooked impact of less severe physical violence on antenatal care visits: Findings from South Asia. Journal of Global Health 2023; 13. DOI:10.7189/jogh.13.04155.

36 Sripad P, Warren CE, Hindin MJ, Karra M. Assessing the role of women’s autonomy and acceptability of intimate-partner violence in maternal health-care utilization in 63 low- and middle-income countries. International Journal of Epidemiology 2019; 48: 1580–92.

37 Wilson-Williams L, Stephenson R, Juvekar S, Andes K. Domestic Violence and Contraceptive Use in a Rural Indian Village. Violence Against Women 2008; 14: 1181–98.

38 Bawuah A, Baatiema L, Sarfo M, Appiah F, Yaya S. The shadow of violence: How intimate partner violence shapes contraceptive and maternal health service use across 25 African countries. PLOS Global Public Health 2025; 5: e0005470.

39 Kaye DK. Community Perceptions and Experiences of Domestic Violence and Induced Abortion in Wakiso District, Uganda. Qualitative Health Research 2006; 16: 1120–8.

40 Pearson E, Andersen KL, Biswas K, Chowdhury R, Sherman SG, Decker MR. Intimate partner violence and constraints to reproductive autonomy and reproductive health among women seeking abortion services in Bangladesh. International Journal of Gynecology & Obstetrics 2016; 136: 290–7.

41 Miller E, Decker MR, McCauley HL, et al. Pregnancy coercion, intimate partner violence and unintended pregnancy. Contraception 2010; 81: 316–22.

42 Moore AM, Frohwirth L, Miller E. Male reproductive control of women who have experienced intimate partner violence in the United States. Social Science & Medicine 2010; 70: 1737–44.

43 Ojha M, Babbar K. Power to choose? Examining the link between contraceptive use decision and domestic violence. Economics & Human Biology 2024; 55: 101416.

44 Sutherland MA. Implications for Violence in Adolescent Dating Experiences. *Journal of Obstetric*, Gynecologic & Neonatal Nursing 2011; 40: 225–34.

45 Thiel de Bocanegra H, Rostovtseva DP, Khera S, Godhwani N. Birth Control Sabotage and Forced Sex: Experiences Reported by Women in Domestic Violence Shelters. Violence Against Women 2010; 16: 601–12.

46 WalterMaurer EM, Ortega CA, McNutt L-A. Issues in Estimating the Prevalence of Intimate Partner Violence: Assessing the Impact of Abuse Status on Participation Bias. Journal of Interpersonal Violence 2003; 18: 959–74.

47 McNutt L-A. Intimate Partner Violence Prevalence Estimation using Telephone Surveys: Understanding the Effect of Nonresponse Bias. American Journal of Epidemiology 2000; 152: 438–41.

48 Gunarathne L, Bhowmik J, Apputhurai P, Nedeljkovic M. Factors and consequences associated with intimate partner violence against women in low- and middle-income countries: A systematic review. PLOS ONE 2023; 18: e0293295.

49 Vieira EM, Perdona GSC, Santos MA dos. Fatores associados à violência física por parceiro íntimo em usuárias de serviços de saúde. Revista de Saúde Pública 2011; 45: 730–7.

50 Amir-ud-Din R, Idrees R, Farooqui J, Abbasi AS. Exploring spousal disparities: Age, earnings, and education as predictors of intimate partner violence in 29 developing countries. Women’s Health 2024; 20. DOI:10.1177/17455057241310289.

51 Asadi S, Mohammad-Alizadeh-Charandabi S, Yavarikia P, Mirghafourvand M. Socio-Demographic Predictors of Intimate Partner Violence in a Population Sample of Iranian Women. Shiraz E-Medical Journal 2018; In Press. DOI:10.5812/semj.69735.

52 Hidrobo M, Fernald L. Cash transfers and domestic violence. Journal of Health Economics 2013; 32: 304–19.

53 Bonomi AE, Trabert B, Anderson ML, Kernic MA, Holt VL. Intimate Partner Violence and Neighborhood Income: A Longitudinal Analysis. Violence Against Women 2014; 20: 42–58.

54 Ahmad J, Khan N, Mozumdar A. Spousal Violence Against Women in India: A Social–Ecological Analysis Using Data From the National Family Health Survey 2015 to 2016. Journal of Interpersonal Violence 2019; 36: 10147–81.

55 Heise LL. Violence Against Women: An Integrated, Ecological Framework. Violence Against Women 1998; 4: 262–90.

56 Sardinha L, Yüksel-Kaptanoğlu I, Maheu-Giroux M, García-Moreno C. Intimate partner violence against adolescent girls: regional and national prevalence estimates and associated country-level factors. The Lancet Child & Adolescent Health 2024; 8: 636–46.

57 Zhao S, Liu S, Gao J, et al. Prevalence of co-occurring forms of intimate partner violence against women aged 15–49 and the role of education-related inequalities: analysis of Demographic and Health Surveys across 49 low-income and middle-income countries. eClinicalMedicine 2025; 82: 103150.

58 Decker MR, Benning L, Weber KM, et al. Physical and Sexual Violence Predictors. American Journal of Preventive Medicine 2016; 51: 731–42.

59 Chakrabarty M, Singh A, Mohan D, Singh S. Understanding the association between intimate partner violence and sexually transmitted infections among women in India: a propensity score matching approach. Sexually Transmitted Infections 2024; 100: 143–9.

60 OJHAS: 2011-3-3. Gaikwad V, Madhukumar S, Sudeepa D. An epidemiological Study of Domestic Violence Against Women and its Association with Sexually Transmitted Infections in Bangalore Rural. http://www.ojhas.org/issue39/2011-3-3.htm (accessed Nov 12, 2026).

61 Grose RG, Grabe S. The Explanatory Role of Relationship Power and Control in Domestic Violence Against Women in Nicaragua: A Feminist Psychology Analysis. Violence Against Women 2014; 20: 972–93.

62 Fay KE, Onwuzurike C, Finley A, Miller E. Integrating anti-violence efforts into sexual and reproductive health: Reproductive coercion as a case example. Contraception 2022; 115: 75–9.

63 Lévesque S, Rousseau C, Boulebsol C, et al. Interpersonal, community, and societal dimensions of reproductive coercion: a sequential multimethod study of victim-survivors. Reproductive Health 2025; 22. DOI:10.1186/s12978-025-02176-x.

